# Accuracy of Attestation Among Mohs and Dermatologic Surgery Diplomates

**DOI:** 10.1101/2022.02.20.22271258

**Authors:** Clifford S. Perlis, Roy H. Perlis

## Abstract

Self-regulation is a key tenet of professionalism. We sought to assess the accuracy of self-attestation with respect to the administration of a newly defined field of board sub-specialization. One qualifying pathway to take the written examination for certification in Mohs and Dermatologic Surgery (MDS) is to self-attest to an active practice of Mohs Surgery. Utilizing publicly available data, we find that greater than 6% (111) of new diplomates did not complete an ACGME-approved fellowship or charge CMS in 2019 for greater than one Mohs case per week. In addition, 72 individuals did not complete a fellowship or charge CMS in 2019 for a single Mohs case. This discrepancy between the test qualifications and practice patterns evident from CMS data suggests concern for the reliability of self-regulation in this setting.

## Introduction

The American Board of Dermatology (ABD) created a sub-specialty certification for Micrographic Dermatologic Surgery (MDS) and announced new diplomates on December 14, 2021. Initial MDS certification required successful completion of an exam given in October 2021, as well as a current medical license, primary ABD certification in dermatology, and current in Maintenance of Certification (MOC). In addition, candidates had to demonstrate experience in the subspecialty through completion of an ACGME-accredited fellowship or “attesting to active practice of micrographic surgery (1).” The ABD did not define “active practice” or require documentation of such. We sought to assess the effectiveness of the self-regulatory process.

## Methods

We assessed how many MDS diplomates, as listed on the ABD website (2), actively practice Mohs surgery using the most recent (2019) available CMS billing data (3). We defined active practice of Mohs surgery a priori as performing an average of one case per week, or 50 cases for the year. As some diplomates may perform Mohs surgery, but may not have billed CMS in 2019, we included only individuals who had at least one CMS charge for either CPT 17000 or 11102 (destruction pre-malignant lesion and shave biopsy).

## Results

Of 1716 MDS diplomates, we eliminated physicians who could not be matched to an NPI number (335), had multiple NPI numbers (36), or lacked CMS charges for either CPT 11102 or 17000 (63). The remaining 1282 individuals (74.7%) were stratified based on the number of times they billed for Mohs surgery (CPT 17311 or 17313). We identified 1109 individuals who charged for greater than 50 cases.

Of the remainder, we excluded 48 individuals who were listed in the ACMS 2021-2022 membership directory as well as an additional 14 individuals who listed completion of an ACGME-accredited fellowship on their own websites, because completion of an ACGME-accredited MDS fellowship also qualifies individuals to take the examination. (That is, these individuals would be eligible regardless of clinical activity). This yielded 111 physicians who achieved MDS board certification without completing a fellowship or averaging more than one Mohs case a week. Among them, 72 did not bill CMS for a single Mohs procedure in 2019, despite billing for other charges.

## Discussion

Study limitations include: some diplomates may actively practice Mohs surgery but not bill CMS, individuals may disagree with our definition for active practice of Mohs surgery, and some diplomates may have completed fellowships that we were unable to identify, as the ACGME does not maintain a list of fellows [4].

Nonetheless, our conservative assumptions may lead us to underestimate the number of individuals who took the MDS exam without actively practicing Mohs surgery. Over 21% of diplomates (371) were eliminated from analysis because we were unable to match each physician to a single NPI number.

Professional organizations granting board certification protect the public by identifying doctors who have achieved certain levels of expertise. Our results indicate that at least 6% of new diplomates did not charge CMS for more than 50 Mohs cases in 2019, including 72 with no record of charging CMS for a single Mohs case in 2019 despite billing for other codes. This discrepancy between the test qualifications and practice patterns evident from CMS data suggests concern for the reliability of self-regulation by attesting physicians and the ABD.

## Data Availability

All data produced in the present work are contained in the manuscript.

